# Examining outdoor play associations in Canadian early learning and child care centres: Cross-sectional insights from the Measuring Early Childhood Outside survey

**DOI:** 10.1101/2025.08.13.25333648

**Authors:** Rachel Ramsden, Barry Forer, Hebah Hussaina, Christina Han, Caroline Bouchard, Jeff Crane, Megan McPhee, Michal Perlman, Mariana Brussoni

**Affiliations:** School of Population and Public Health, University of British Columbia, Vancouver, BC, Canada; British Columbia Children’s Hospital Research Institute, Vancouver, BC, Canada; Human Early Learning Partnership, University of British Columbia, Vancouver, BC, Canada; Department of Pediatrics, University of British Columbia, Vancouver, BC, Canada; Faculté des sciences de l’éducation, Université Laval, Québec, Canada; Faculty of Education, Memorial University of Newfoundland, St. John’s, Newfoundland, Canada; Early Childhood Development Association of PEI, Charlottetown, Prince Edward Island, Canada; Department of Human Development and Applied Psychology, University of Toronto, Ontario, Canada

**Keywords:** child care, national survey, early childhood education, playground, outdoor play space

## Abstract

Canada lacks national data on the current provision of outdoor play (OP) in Early Learning and Child Care (ELCC) programs. In this study, we report results of the Measuring Early Childhood Outside (MECO) national survey to fill this gap and examine the factors that are associated with children’s OP and risky play in ELCC programs. Respondents included ELCC centres providing full-day licensed group care (birth to school entry) in Canada. Primary outcomes measured were OP frequency, OP duration and risky play occurrence. Hierarchical multiple regressions were used to examine relationships and interaction effects between the primary outcomes and 14 variables encompassing centre, staff, physical environment and OP provision characteristics, for infant/toddler-aged and preschool-aged programs separately. A total of 1,187 ELCC centres responded to the MECO survey (9.8% response rate), of which 67.2% were non-profit providers. Most centres went outdoors every day, regardless of the season, though they spent less time outdoors in the winter than in the summer. Risky play was limited, with play at heights being the most common, and use of fire the least common. Variables that emerged as positively associated with most outcomes across programs related to training of centre directors and educators, giving children the autonomy to make decisions about going outdoors, providing all-weather gear, including diverse affordances (loose parts, gardening elements, fixed equipment), having outdoor spaces larger than required by licensing requirements, and the use of off-site spaces. Information about the current state of OP in ELCC centres is important at a time of considerable expansion in the sector, helping inform evidence-based policy development to enhance OP opportunities across Canada.

## Introduction

### Outdoor Play and Child Development and Well-Being

Research outlines the profound benefits of outdoor play (OP) to multiple aspects of children’s development, health and well-being (1–3). Children are naturally driven by curiosity, and outdoor environments are the ideal settings for them to explore, engage their senses, and learn through hands-on experiences (3,4). OP nurtures essential skills that shape a child’s development and sense of self (3). It invites open-ended interactions, spontaneity, risk-taking, exploration, discovery, and connection with nature. In addition, OP cultivates social and emotional growth by helping children build social skills, learn cooperation, and resolve conflicts effectively (5,6).

OP differs from indoor play by offering its own distinct benefits, including being more inclined towards dynamic and vigorous physical activities (7,8). Engagement in OP supports children’s direct contact with nature, allowing them to physically interact with plants and receive rich sensory stimulation that activates multiple senses. Natural spaces can inspire deeper engagement and a profound sense of wonder in children (9). Additionally, outdoor environments support visual health by enabling children to focus on distant objects (10), and they provide natural exposure to sunlight, which facilitates the body’s production of vitamin D — benefits that are typically limited or unavailable in indoor settings (11).

OP is also more likely to consist of risky play, a thrilling form of physical play involving uncertainty and the potential for injury (12). Risky play includes: 1) Play with great heights; 2) Play with high speed; 3) Play with dangerous tools; 4) Play near dangerous elements; 5) Rough-and-tumble play; 6) Play where children go exploring alone; 7) Play with impact; and 8) Vicarious play (13,14). Activities such as climbing, running, exploring independently, and rough-and-tumble play can contribute to physical activity and motor skill development, and cognitive development (12,15). Risky play allows children to test their limits, experience feelings of exhilaration and fear, and develop critical skills (6,15,16). Children assess risks, make plans and decisions, and solve problems. For example, deciding how high to climb or how fast to swing involves complex cognitive calculations related to capabilities, conditions and preferences (17). Further, engaging in manageable levels of fear and stress helps children regulate emotions, such as fear and excitement, while building resilience and self-confidence (18). By testing their limits and understanding their abilities in controlled settings, children gain a sense of mastery, modulate their emotions, and reduce fear responses in other situations – fostering essential life skills and emotional growth (16,18,19).

Three key ingredients – time, space and freedom – have been identified as necessary for OP opportunities (6). *Time* involves making play a daily priority, as important as sleep or other basic functions. *Space* relates to ensuring that children have easy access to stimulating play spaces replete with diverse affordances for play. *Freedom* involves allowing children to play in the ways they choose, including taking risks that naturally emerge during play. Despite the numerous benefits, there is an evident decline in OP (20), especially in Western societies, which has been attributed to factors related to each of these key ingredients (time, space and freedom) (6,21–24). The time dedicated to OP has shrunk as structured academic activities indoors have been prioritized, and screens have become ever more enticing (20). Outdoor spaces for play have become less accessible and appealing with increasing urbanization and traffic (25), and an emphasis on safety has led to play spaces with limited challenges (26,27). Children’s freedom has been curtailed by risk-averse parents, educators and play providers concerned about injury and liability (21,28).

### OP in Early Learning and Child Care Programs

In efforts to reverse the declining trend of OP, it becomes important to consider the places where children already spend their days and the opportunities for increasing time, space and freedom in these settings. Early learning and child care (ELCC) centres are important environments to examine with children under the age of 6 in many countries spending most of their waking hours in child care (29). In Canada, 56% of children aged 0 to 5 years are in some form of child care, an increase from prior years (30).

OP in ELCC programs has been influenced by the broader societal trends and the key ingredients of time, space and freedom have been compromised. These factors can be mapped onto the various levels of Ecological Systems Theory (31,32). At the societal (macrosystem) level, social and cultural norms that have devalued the importance of OP have extended their influence into the ELCC sector, limiting *time* for play as ELCC environments also face the pressures for, and the perception that time is better spent in academic preparation, and have limited understanding of the developmental, well-being and academic benefits of play outdoors (33). ELCC professionals may not have the background to plan for meaningful outdoor experiences, may have negative attitudes toward OP, or face competing interests from scheduling and family influences, further reducing time allocated outdoors in ELCC centres (32,34).

Further, rapid urbanization at the societal level has put pressure on the *spaces* available for OP, with outdoor areas often the last to be prioritized in ELCC centres (35–37). Thus, at the centre (microsystems) level, this means that many centres entirely lack outdoor spaces and in others, these spaces are often limited in size, poorly designed, and lack the natural elements that foster creativity, exploration, and diverse forms of play (38,39).

At the organizational and regulatory (exosystem) level, licensing and other policies have restricted *freedom* for OP (40). Regulations and licensing requirements, while essential for safety, can have unintended consequences, particularly when they are interpreted in restrictive ways (32). Playground safety standards can be excessively rigid and do not consider the local context and capabilities of the children playing in those spaces, leading to overly cautious designs that remove essential elements of challenge and exploration from children’s play (32,41). Additionally, the mesosystem level interaction between environments means that at the centre (microsystems) level, safety concerns and risk aversion among families and educators, compounded by educators’ fears of liability, can make it difficult to balance safety with the need for children to experience challenge and risk-taking (32,42). Furthermore, educators’ perspectives and training play an important role, as some may not see the value of spending time outdoors or lack confidence in supervising OP or advocating for risk-taking activities, leading them to opt for “safer,” indoor alternatives (32,40,43). The balance between educator restriction, guidance, and scaffolding plays a pivotal role in shaping the degree of freedom afforded to children during OP experiences (44).

Despite the challenges, there are promising developments emerging within Canadian ELCC programs. Policymakers and practitioners are increasingly recognizing the need for a more balanced approach to facilitating risk and challenge in OP (40). The importance of OP is gaining traction, with an emphasis on providing children with diverse play affordances, including risky play (45). By offering consistent and extended access to OP throughout the day and across various seasons, ELCC programs have the unique potential to make OP a fundamental and routine component of children’s daily lives (40). Enhancing OP opportunities in ELCC programs plays a critical role in supporting children’s health, well-being and development.

### Current State of OP Provision and Policies in Early Learning and Child Care in Canada

The ELCC landscape in Canada is rapidly shifting and it is an opportune time to examine the current status of OP, as well as consider mechanisms for increasing access. The federal government of Canada launched two major initiatives: 1) the Multilateral Early Learning and Child Care Framework (MELCCF); and 2) the Canada-wide Early Learning and Child Care (CWELCC) plan (46,47). Introduced in 2017, the MELCCF, guided by principles of quality, accessibility, affordability, flexibility, and inclusivity, aims to enhance ELCC services and their workforce, programs, and learning environments – particularly for children from Indigenous, rural and diverse needs backgrounds. Building on the MELCCF’s foundation, the CWELCC was introduced in 2021 to provide funding to provinces and territories to support the expansion of child care services, enhance quality, and reduce fees for families, with the goal of lowering the average cost of child care to $10 per day by 2026. These efforts represent significant strides towards addressing the child care needs of Canadian families; however, ongoing debates continue over the most effective approach for advancing as a nation in addressing the sustainability of funding mechanisms and the need for further policy reforms to ensure equitable access to high-quality and accessible child care for all children (37,48). Neither of these initiatives addresses children’s OP provision, and the rapid efforts to expand ELCC services can limit OP by exacerbating pressures on available outdoor spaces and compromising opportunities for educators to receive proper training on OP (49).

While the federal government in Canada plays a role in funding and setting broad policy frameworks for ELCC, the direct responsibility for designing, regulating, and delivering ELCC services, including those related to OP, rests with individual provinces and territories (50). As a result, the current state of OP in ELCC in Canada is characterized by a complex and fragmented landscape, marked by regional inconsistencies in policies, regulations, and practices (51). That said, provincial and territorial jurisdictions vary widely in their curricular emphasis on OP, with some program guides making minimal mention of it (50). This disparity extends to the regulation of OP practices, with wide variations in required daily minutes, minimum outdoor space requirements, and safety standards (50). For example, the minimum outdoor space requirement per child for infants differs widely between Alberta (2 m^2^ per child) and Prince Edward Island (7 m^2^ per child). Some jurisdictions do not require dedicated OP space connected to the ELCC centre, while others only require outdoor space for a percentage of participating children (50). Further, licensing requirements vary regarding mandatory minimum spent outdoors, with some jurisdictions having no minimum requirement, and others, ranging from 60 minutes (British Columbia, New Brunswick,) to 120 minutes daily (Ontario) (50). Such inconsistencies contribute to a patchwork system where children’s access to quality OP experiences is heavily dependent on their geographic location and the corresponding regulations.

The overall picture of OP in Canadian ELCC is one of ongoing evolution and adaptation. While significant challenges remain, an increased awareness of the importance of children’s OP offers hope for a future where Canadian children have more equitable access to enriching and developmentally beneficial OP experiences. To support and enhance OP in Canadian ELCC settings, the existing political framework governing ELCC provision must prioritize OP opportunities within the federal, provincial and regional initiatives and regulations.

### Measuring Early Childhood Outside (MECO) Pan-Canadian Survey

The Measuring Early Childhood Outside (MECO) pan-Canadian survey was designed to capture the current provision of OP in ELCC programs in Canada, including the frequency and duration of OP, and permitted risky play activities. This national survey also strived to examine and measure theoretically relevant variables that are known to influence children’s OP behaviour in ELCC environments. Existing evidence outlines centre-level predictors (i.e., auspice, geographic location and age groups served) (37), staff-related predictors (i.e., professional development, training and educator tenure)(32,34), physical environment-level predictors (i.e., OP play space size, loose parts, gardening areas and equipment types) (52), and provision-related predictors (i.e., all-weather gear and child autonomy) (32,43) of children’s OP. Although these predictors shape opportunities for children’s outdoor and risky play, their individual and combined influence on the current state of play provision in ELCC programs in Canada remains undiscovered. The MECO survey aimed to explore this relationship through the framework of time, space, and freedom (Figure 1).

**Figure 1:**
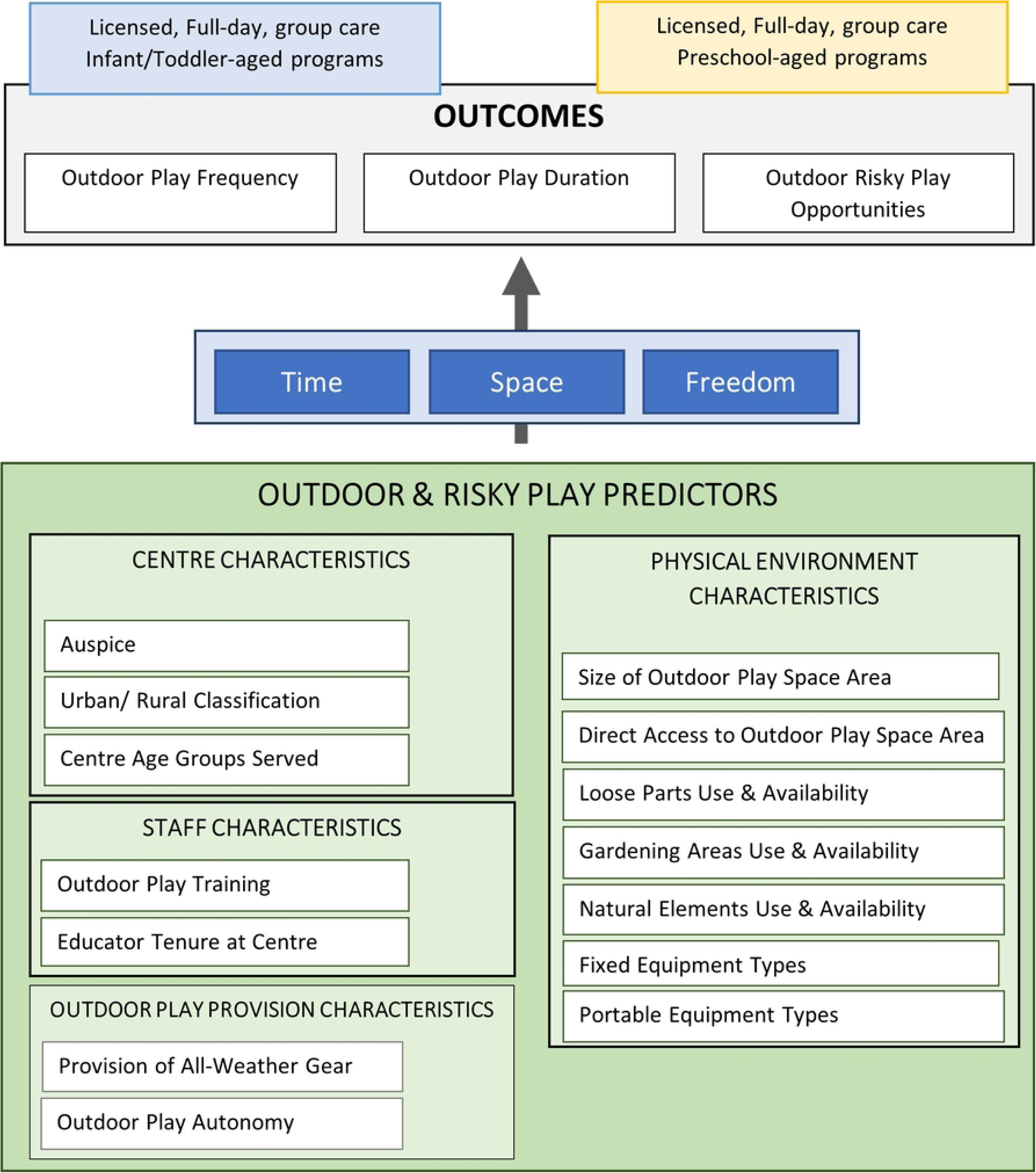
A conceptual model of how centre, staff, physical environment and additional characteristics influence outdoor play and risky play outcomes through the constructs of time, space and freedom.

Despite the growing recognition of the importance of OP in ELCC settings, we are aware of no Canadian data reporting on OP provision in these settings to-date. National data on the current status of OP in ELCC settings is imperative to identify gaps in the ELCC system, plan effective policies and strategies for improving OP opportunities, and contribute important knowledge to the research landscape. This paper aims to provide a cross-sectional overview on the current status of children’s outdoor and risky play participation in Canada. Secondly, this paper assesses the association between important predictors of children’s OP and children’s outdoor and risky play outcomes. Investigating important predictors of children’s OP supports an enhanced understanding of associations with children’s OP, but does not make any causal claims on relationships.

## Methods

### Setting and Inclusion Criteria

The target population for the MECO survey was ELCC centres in Canada providing full-day, group care to children before entry to primary school (birth to school entry age). This survey excluded nursery schools, school-age programs, preschools and other part-day programs, licensed family or in-home child care, and unlicensed programs due to the inconsistencies in outdoor programming and space-related regulatory standards within provincial and territorial policies. Limiting the survey to full-day, licensed ELCC group care centres allowed for a shared context for more detailed questions that were applicable to all respondents. Furthermore full-day group care ELCC centres deliver care to large groups of children and are expected to expand as an important child care option for Canadian families.

### Data Collection Procedure

#### Survey Development and Pilot Testing

The MECO survey was developed with input from a literature review and the advisory committee. A literature review was conducted to find existing survey research that could inform survey development and survey questions previously used to measure children’s OP provision. The study was guided by an advisory committee that included individuals from ELCC organizations and academic institutions across Canada. The committee set priorities for the survey, reviewed survey drafts, recommended recruitment strategies, and assisted with interpreting the results. The MECO survey was developed in English and subsequently translated into French. The English and French versions of the survey were hosted on REDCap (53), a secure, password-protected data collection platform hosted through the [redacted for review]. To ensure clarity and usability, the survey was pilot-tested with 9 ELCC providers (5 English-speaking and 4 French-speaking) through cognitive interviews. These interviews helped evaluate the content and comprehension of survey questions. Based on feedback from these sessions, the survey was iteratively revised. The final survey included eligibility screening questions and questions relating to the ELCC centre characteristics (e.g., auspice, staffing, size), program type, characteristics of the children attending the program, days and hours spent in OP (in summer and winter), factors influencing OP time (e.g. licensing regulations, weather, etc.), characteristics of the outdoor space (e.g., size, access, loose and fixed parts), permitted outdoor risky play activities, other outdoor locations, provision of all-weather gear, and staff training and experience. Respondents were also invited to upload photos of their outdoor space, and share any other relevant information. The administered version of the MECO survey is provided in S1 Appendix.

#### Survey Distribution and Promotion

The [redacted for review] provided approval for the research (#H22-00210). To distribute the survey, a comprehensive list of eligible ELCC programs across Canada was compiled from public databases, government contacts, and regional organizations. Where direct email contact information could not be obtained, outreach was done through trusted provincial or regional organizations. The survey was distributed via email to all provinces and territories, except Alberta and Ontario. In these two provinces, alternative strategies were used (e.g., social media promotion, collaboration with local advocacy groups, and posting in community newsletters) due to the lack of publicly available or easily reachable contacts. Additionally, no outreach was done to ELCCs in the Northwest Territories (NWT) as we were unable to obtain a research license to become eligible to collect data in the NWT. Social media promotion was initially employed but halted in August 2023 due to an influx of automated (i.e., ‘bot’) responses.

Ongoing promotion included reminder emails sent to regional, provincial, territorial, and national organizations, and through newsletters such as the OP Canada Newsletter and e-blasts from the Canadian Child Care Federation. The survey was also promoted at the Breath of Fresh Air Conference in September 2023.

#### Survey Administration

The MECO survey collected responses between June 23, 2023 and November 6, 2023 in both English and French. We requested that the survey be completed by only one respondent for each ELCC centre and required that the respondent was the individual most responsible for the day-to-day operations of the centre. If respondents were responsible for the operation of multiple centres in separate physical addresses, they were instructed to complete a separate survey for each centre. Respondents who completed the survey were incentivized with a $25 gift card. To ensure data quality, enhanced security features were added to the survey (reCAPTCHA technology), email authentication, and ‘honey pot’ style verification questions tailored to ELCC-specific knowledge. Further, the research team validated responses by screening for automated bot submissions and duplicate responses. Examples of screening methods to eliminate automatic bot responses include examining overall survey response time (<3 minutes was determined as not possible for verified responses), reviewing responses to ‘honey pot’ questions, and examining open-text and photo submissions for artificially generated or stock responses. After the survey closed, verified responses were de-identified, assigned unique IDs, and exported from the REDCap platform for analysis.

### Survey Response Rates and Weighting

A total of 1,187 respondents completed the MECO survey. Table 1 shows, for each province and the territories, the number of full-day group ELCC centres in the population, the number of responding centres, and the corresponding response rate. The total population was derived from the number of full-day child care centres published in the Early Childhood Education and Care in Canada 2021 report (54). Overall, 9.8% of eligible ELCC centres responded to the survey. Four provinces had response rates above 20%, and five provinces had response rates below 10%. Due to low response rates, and ineligible participation from NWT, responses from Nunavut and Yukon territories were combined to make up the ‘territories’ jurisdiction. To rebalance these disproportionalities, responses were weighted while keeping the total number equal to the number of survey responses. As a result of weighting, the proportion of responses in each jurisdiction was restored to the population proportions, allowing a truer estimate of results for Canada as a whole.

**Table 1.** MECO survey ELCC centre response rates, Canada and jurisdictions.

| **Jurisdiction** | **Population**<br>**(Number of**<br>**Centres in Canada)** | **Proportion of**<br>**Total Canadian**<br>**Population (%)** | **Sample (Number**<br>**of Centres**<br>**Responding)** | **Proportion of**<br>**Total Survey**<br>**Sample (%)** | **Survey**<br>**Response**<br>**Rate (%)** |
| --- | --- | --- | --- | --- | --- |
| **Canada** | **12,076** | **100.0** | **1,187** | **100.0** | **9.8** |
| British Columbia | 2,117 | 17.5 | 466 | 39.3 | 22.0 |
| Alberta | 1,113 | 9.2 | 66 | 5.6 | 5.9 |
| Saskatchewan | 346 | 2.9 | 80 | 6.7 | 23.1 |
| Manitoba | 612 | 5.1 | 128 | 10.8 | 20.9 |
| Ontario | 3,312 | 27.4 | 154 | 13.0 | 4.6 |
| Quebec | 3,544 | 29.3 | 127 | 10.7 | 3.6 |
| New Brunswick | 422 | 3.5 | 40 | 0.3 | 9.5 |
| Nova Scotia | 271 | 2.2 | 100 | 8.4 | 36.9 |
| Prince Edward Island | 78 | 0.6 | 8 | 0.7 | 10.3 |
| Newfoundland and Labrador | 149 | 1.2 | 14 | 1.2 | 9.4 |
| Territories<sup>a</sup> | 112 | 0.9 | 4 | 0.3 | 3.5 |

<sup>a</sup> Population includes Nunavut, NWT and Yukon; Sample includes only Nunavut and Yukon

### Analysis

#### Outcome Variables for OP Duration, OP Frequency, and Risky Play

All the outcome variables described below were captured separately for infant/toddler (I/T) programs and preschool-aged (PS) programs – even if the centre served children in both age groups. Respondents self-identified the age categories (I/T and/or PS) their program served, therefore the specific ages of each age category may differ by jurisdiction and as defined by regional licensing regulations. Seasonality was considered for the outcome variables OP duration (i.e., typical number of hours per day) and frequency (i.e., typical number of days per week), with responses captured separately for each season. For these analyses, responses from winter and summer were only considered to understand outcome differences between contrasting seasons. Altogether, eight outcomes were modeled: four for OP duration, two for frequency, and two for risky play (see below). Frequency of OP in the summer months was not modeled, as over 95% of centres reported daily frequency, leaving little remaining variability for modeling.

OP frequency (“In a typical week in each season, how many days are [the children] taken outdoors?” with response options from “0 days” to “5 or more days”) was captured for each of the four combinations of season and age group of children. Responses were transformed into a quantitative value between 0 and 5, with the lowest response category (“0 days”) assigned a score of 0 and the highest category (“5 or more days”) assigned a score of 5. OP duration (“On a typical day in each season, approximately how many hours does each [child] spend outdoors”? with response options ranging from “none” to “5 or more hours”) was captured for each of the four combinations of season and age group of children. Respondents answered based on a six-point scale, with a range of hours associated with each point. Duration responses were transformed to the mid-point of the range. For example, if a respondent chose “3 to 4 hours,” their assigned numeric score was 3.5.

Risky play was assessed through four categories of activities – play at heights, play with tools, use fire, and rough-and-tumble play – in alignment with four of Sandseter’s risky play categories (13,14). Additional categories of risky play (play with high speed, play where children go exploring alone, play with impact, and vicarious play) were not measured in the MECO survey. For each of these activities, respondents were asked “How often do [children] at your program have opportunities to do the following activities outdoors?” For each item, respondents could select among five “how often” categories: 1) “not allowed”, 2) “never”, 3) “sometimes”, 4) “often” and 5) “always.” For the purpose of our analyses, “not allowed” and “never” were collapsed into one category. Assuming roughly equal intervals between the categories, an overall risky play score was calculated as the sum of the item scores, resulting in a total score ranging from 0 to 12. The derived risky play score was used for the risky play outcome in our analyses. Unlike the OP duration and OP frequency outcomes, seasonality was not captured for the risky play outcome.

#### Explanatory Variables

Fourteen variables were selected a priori for inclusion in multivariate regression modelling and grouped into four categories: 1) centre-level organizational characteristics; 2) staff-related characteristics; 3) physical environment characteristics; and 4) OP provision characteristics [37]. An overview of all included explanatory variables are provided in Table 2 and further detail on how these variables were derived from the MECO survey is outlined in S2 Appendix.

**Table 2:** List of 14 explanatory variables included in the analysis, summarized by category and variable levels.

| **Explanatory Variable** | **Category** | **Variable Levels** |
| --- | --- | --- |
| Auspice | Centre-level | 2 |
| Urban/Rural Classification | Centre-level | 2 |
| Centre Age Groups | Centre-level | 2 |
| OP Training | Staff-related | 4 |
| Educator Tenure at Centre | Staff-related | 3 |
| Size of OP Area | Physical Environment | 4 |
| Access to OP Area | Physical Environment | 2 |
| Count of Loose Parts | Physical Environment | 9 |
| Count of Gardening Areas | Physical Environment | 5 |
| Count of Natural Elements | Physical Environment | 7 |
| Count of Fixed Equipment Types | Physical Environment | 9 |
| Count of Portable Equipment Types | Physical Environment | 11 |
| Provision of All-Weather Gear | OP Provision | 3 |
| Children's OP Autonomy | OP Provision | 3 |

### Analytical Plan

Descriptive results are presented related to the number of respondents (N) for each variable. For each of the eight outcomes (summer OP duration, winter OP duration, winter OP frequency and risky play occurrence, for I/T and PS programs), a hierarchical multiple regression strategy was employed. Multiple models assessed the influence of each group of predictors on the outcomes, with the final model including all explanatory variables. A secondary model included interactions between staff training and each of three outdoor characteristics – loose parts, fixed equipment, and child autonomy (S3 Appendix). Interaction terms were included based on prior knowledge of the influence staff training has on additional predictors of children’s OP, such as loose parts inclusion (Spencer et al., 2019). As suggested by Sawilowsky (2009), betas of .200 or greater in the regression models were considered to reach the level of at least a small effect size. The full results of these eight hierarchical regression models are located in S3 Appendix.

## Results

### Descriptive Results for Outcome Variables

Weighted descriptive results for the three outcome variables modeled in this analysis are provided in Table 3. The results for these outcomes are separated for I/T and PS programs, and by summer and winter seasons for OP frequency and duration outcomes. Approximately 75% of responding centres offered both I/T– and PS-aged programs, therefore results separated by program type (I/T, PS) contain sample sizes larger than the 1,187 centres in the sample. For both I/T and PS programs, the mean OP frequency in summer is approximately five days per week (i.e., every day), therefore these outcomes were not modelled. I/T programs reported a mean OP frequency of 4.5 days per week outdoors in the winter, whereas PS programs reported a mean OP frequency of 2.9 days per week. To further understand the low reported OP frequency by PS programs, Table 4 provides OP frequency descriptive results by whether the centre serves only PS children or both age groups. The results show that the lower winter frequency of OP for PS children only applies when centres offer programs for both age groups. Both I/T and PS programs reported longer OP duration in the summer than in the winter months. The mean risky play score was 2.5 for I/T programs, and 3.8 for PS programs. Of the four activities, the one contributing the most to this difference between the age groups was the frequency of play at heights.

**Table 3.** Weighted descriptive results for all outcome variables, by program type (infant/toddler & preschool-aged).

| Program Type | Outcome Variable | Median | Mean | SD | Range | N |
| --- | --- | --- | --- | --- | --- | --- |
| Infant/<br>Toddler | OP Frequency, Summer (days/ week) | 5.0 | 4.93 | 0.40 | 0 to 5 | 964 |
|  | OP Frequency, Winter (days/ week) | 5.0 | 4.51 | 0.93 | 0 to 5 | 967 |
|  | OP Duration, Summer (hours/ day) | 2.5 | 2.87 | 1.19 | 0 to 5 | 967 |
|  | OP Duration, Winter (hours/ day) | 1.5 | 1.64 | 1.01 | 0 to 5 | 969 |
|  | Play at Heights | 1.0 | 1.03 | 1.08 | 0 to 3 | 962 |
|  | Play with Tools | 0.0 | 0.34 | 0.69 | 0 to 3 | 961 |
|  | Use Fire | 0.0 | 0.07 | 0.35 | 0 to 3 | 963 |
|  | Rough-and-tumble Play | 1.0 | 1.10 | 1.02 | 0 to 3 | 964 |
|  | Risky Play (total score) | 2.0 | 2.52 | 2.24 | 0 to 12 | 967 |
| Preschool-aged | OP Frequency, Summer (days/ week) | 5.0 | 4.90 | 0.53 | 0 to 5 | 1,080 |
|  | OP Frequency, Winter (days/ week) | 3.0 | 2.93 | 1.23 | 0 to 5 | 1,097 |
|  | OP Duration, Summer (hours/ day) | 3.5 | 3.28 | 1.22 | 0 to 5 | 1,092 |
|  | OP Duration, Winter (hours/ day) | 1.5 | 2.04 | 1.03 | 0 to 5 | 1,099 |
|  | Play at Heights | 2.0 | 1.64 | 1.10 | 0 to 3 | 1,100 |
|  | Play with Tools | 0.0 | 0.58 | 0.79 | 0 to 3 | 1,101 |
|  | Use Fire | 0.0 | 0.11 | 0.42 | 0 to 3 | 1,096 |
|  | Rough-and-tumble Play | 1.0 | 1.47 | 1.02 | 0 to 3 | 1,102 |
|  | Risky Play (total score) | 4.0 | 3.79 | 2.36 | 0 to 12 | 1,105 |

**Table 4.** Weighted descriptive results for outdoor play frequency in winter for preschool-aged programs, by age groups served at responding ELCC centre (weighted).

| Age Groups Served | Outcome Variable | Median | Mean | SD | Range | N |
| --- | --- | --- | --- | --- | --- | --- |
| Centre Serves PS Children Only | OP Frequency, Winter (days/week) | 5.0 | 4.47 | 1.02 | 1 to 5 | 211 |
| Centre Serves I/T and PS Children | OP Frequency, Winter (days/week) | 2.0 | 2.56 | 0.96 | 0 to 5 | 886 |

### Descriptive Results for Explanatory Variables

Weighted descriptive results of investigated explanatory variables are summarized in Table 5-7. Table 5 outlines the sample characteristics of respondents related to centre-level and staff-related characteristics. The weighted sample primarily consists of non-profit centres (67.2%), most often offering services to both I/T and PS children (75.4%), and located in urban locations (82.2%). Responding centres most frequently had no director or educators with OP-specific training (41.8%), or both the director and educators had received OP training (30.8%). Educator tenure across the responding centres varied, with the largest percentage of respondents (39.8%) indicating the majority of educators (>50%) had been at their ELCC centre at least five years.

**Table 5:** Weighted descriptive results for centre-level and staff-related characteristics of responding ELCC centres.

| Explanatory variable | Total |
| --- | --- |
| **Centre Characteristic** |  |
| Auspice [n(%)] |  |
| For-Profit | 358 (30.1%) |
| Non-Profit | 798 (67.2%) |
| Indigenous-led Non-Profit<sup>a</sup> | 25 (2.1%) |
| Unknown<sup>b</sup> | 7 (0.5%) |
| Urban/Rural Classification [n(%)] |  |
| Urban | 976 (82.2%) |
| Rural | 211 (17.8%) |
| Centre Age Groups [n(%)] |  |
| I/T Only | 78 (6.6%) |
| PS Only | 214 (18.0%) |
| Both I/T and PS | 895 (75.4%) |
| **Staff-related Characteristic** |  |
| OP Training [n(%)] |  |
| None | 493 (41.8%) |
| Director Only | 211 (17.6%) |
| Educators Only | 111 (9.4%) |
| Educators and Director(s) | 363 (30.8%) |
| Proportion of Staff Educators with Five Years or Longer at the Centre [n(%)] |  |
| Under 25% | 388 (33.4%) |
| 25% to 49.9% | 312 (26.9%) |
| 50% and Over | 462 (39.8%) |

<sup>a</sup> Included in non-profit category for all regression models
<sup>b</sup> Not modelled

Summarized weighted descriptive results related to physical environment and OP provision characteristics are provide in Table 6 (I/T programs) and Table 7 (PS programs). Most respondents reported that children have direct access to the OP areas from the indoor areas (81.9% and 78.5%, respectively), and their OP spaces were larger than required by licensing regulations (70.1% and 68.4%, respectively). The most-provided elements in OP areas for I/T and PS programs were portable equipment and loose parts. PS programs had higher mean scores than I/T programs for counts of loose parts, gardening areas, natural elements and fixed and portable equipment types. For both I/T and PS programs, almost half of respondents reported no provision of all-weather gear to either children or educators (46.0% and 48.5%). Where all-weather gear was provided, it was most often provided to children but not educators. Children in PS programs (17.6%) were more likely than children in I/T programs (14.5%) to have the autonomy to decide when to go to the OP area; however, the majority of programs indicated children’s autonomy was rare or non-existent (43.6% and 51.1%).

**Table 6.**
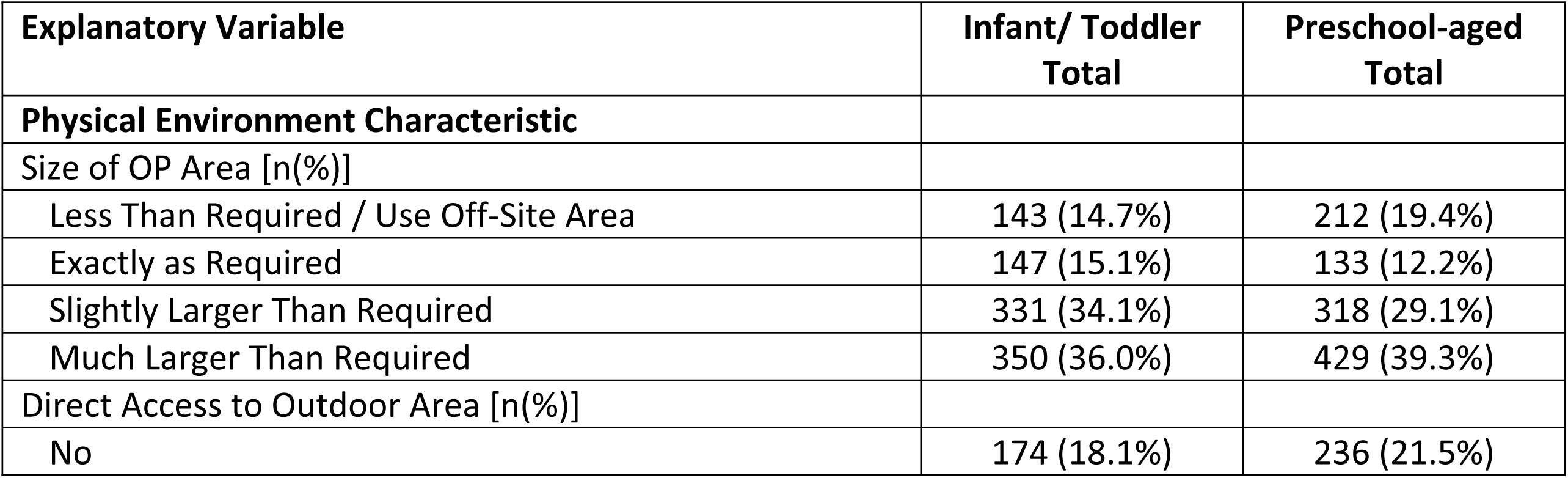
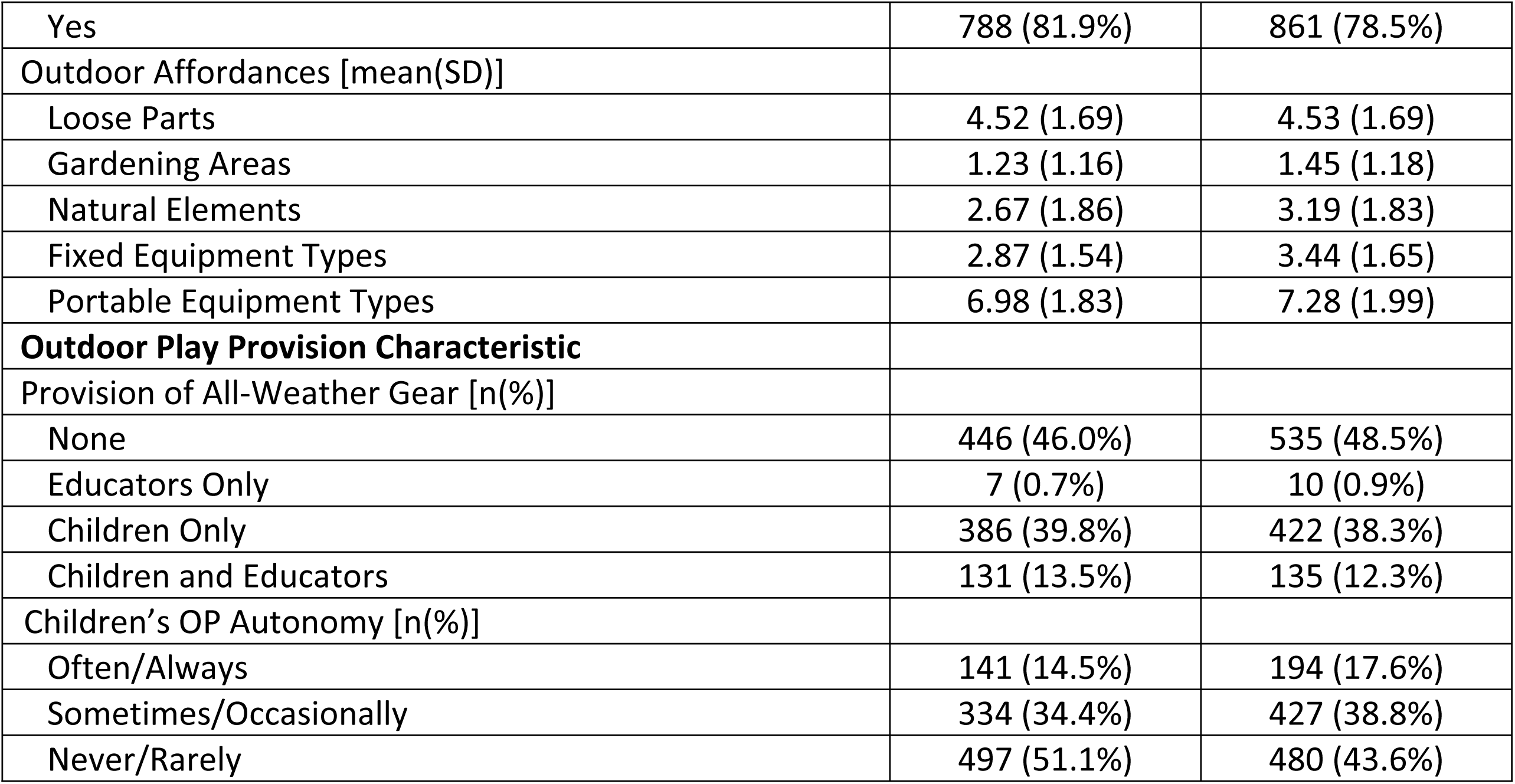
Weighted descriptive results for physical environment and OP provision characteristics of responding ELCC centres, by program type (infant/toddler & preschool-aged).

**Table 7.** Regression model results for ELCC centres serving infant/ toddler programs, all outcomes.

|  |  | All Outcomes: Infant/Toddler Programs, Betas & Significance |  |  |  |
| --- | --- | --- | --- | --- | --- |
|  |  | Duration,<br>Summer | Duration,<br>Winter | Frequency,<br>Winter | Risky Play |
| Auspice | Non-profit<br>vs. For-profit | **-.127\*\*\*** | -.020 | .027 | -.056 |
| Rural/Urban | Rural vs.<br>Urban<br>Postal Code | .017 | -.038 | -.010 | .011 |
| Centre Ages | Both Ages<br>vs. Just I/T | -.062 | **-.075\*** | -.042 | -.042 |
| OP Training | Director<br>Only vs.<br>None | .05 | .044 | -.003 | .011 |
|  | Educator<br>Only vs.<br>None | .05 | .027 | -.038 | .005 |
|  | Director<br>and<br>Educators<br>vs. None | -.008 | .043 | **.125\*\*\*** | **.098\*\*** |
| Staff Tenure | Medium vs.<br>Low | .02 | -.064 | -.013 | -.047 |
|  | High vs.<br>Low | .055 | -.064 | -.047 | **-.096\*\*** |
| OP Area Size | Smaller/Off-Site vs. As<br>required | .013 | -.033 | .028 | .061 |
|  | Larger vs.<br>As required | .019 | .021 | **.155\*\*** | -.060 |
|  | Much larger<br>vs. As<br>required | **.114\*** | -.010 | .086 | .054 |
| Access to OP<br>Area | Direct vs.<br>Not | **.071\*** | -.045 | -.031 | **.056** |
| Affordances | Loose Parts | **.142\*\*\*** | **.153\*\*\*** | **.109\*** | **.164\*\*\*** |
|  | Gardening<br>Elements | **.097\*** | **.118\*\*** | .021 | **.119\*\*** |
|  | Natural<br>Elements | -.041 | .046 | .062 | .008 |
|  | Fixed<br>Equipment | **.153\*\*\*** | -.030 | -.045 | **.138\*\*\*** |
|  | Portable<br>Equipment | -.039 | -.051 | .069 | -.030 |
| All-Weather Gear | Children or Educators vs. Neither | .058 | .036 | .039 | .043 |
|  | Children and Educators vs. Neither | **.076\*** | **.107\*\*** | .061 | **.086\*\*** |
| Child Autonomy | Medium vs. Low | **.128\*\*\*** | .057 | .008 | **.165\*\*\*** |
|  | High vs. Low | **.066\*** | **.090\*\*** | -.008 | **.230\*\*\*** |
|  | Adjusted R<sup>2</sup> | .135 | .076 | .067 | .254 |
|  | N (weighted) | 901 | 903 | 901 | 902 |

\* $p < .05$ , \*\* $p < .01$ , \*\*\* $p < .001$

### Multiple Regression Results Across OP Outcomes

#### Infant/Toddler Programs

To understand the influence of the examined explanatory variables on OP opportunities in I/T programs, a final hierarchal regression model was employed for each outcome variable (Table 7). Type of auspice had a small but significant effect for OP duration, with for-profit centres reporting longer OP duration in comparison to non-profit centres. Winter OP duration for I/T programs was shorter for centres serving both I/T and PS age groups (β=-.075) in comparison to centres serving only I/T aged children. Formal training in OP was associated with increased OP frequency in winter as well as more risky play, but only for centres where both the director and educators had OP training. The composition of educator tenure was only significantly associated with risky play. Although a small effect, higher proportions of long-tenured educators was negatively associated with risky play (β=-.096). Children’s autonomy to go outdoors, both medium (β=.165) and high (β=.230), autonomy, were positively associated with increased risky play, as well as OP duration in both summer (β=.066) and winter (β=.090). The provision of all-weather gear to both children and educators was significantly related to greater duration of OP in both summer (β=.076) and winter (β=.107), and to more risky play (β=.086). Frequency of OP in winter was unrelated to gear provision for educators or children.

Characteristics relating to the physical environment of OP areas were associated with most examined I/T-aged program outcomes. Direct access to the OP area from the indoor program space was positively associated with summer OP duration (β=.071) and risky play (β=.056). Centres with larger OP areas than required by licensing regulations were significantly associated with increased OP frequency in the winter months (β=.155), and much larger OP areas were positively associated with summer OP duration (β=.114). I/T programs that relied on off-site OP areas to meet licensing requirements or had less than required on-site outdoor space were not significantly different than those with the minimum licensed outdoor areas for any of the four I/T outcomes. The count of loose parts was associated with increased OP frequency (β=.109) and OP duration (β=.153) in the winter. Additionally, OP duration in the summer (β=.097) and in the winter (β=.118) were positively associated with the number of gardening elements. The number of fixed equipment affordances within the OP area was significantly associated with I/T summer OP duration (β=.153). Risky play was the only outcome without an association with the number of loose parts, though there was an association with both the number of types of gardening elements (β=.119) and fixed equipment (β=.138).

Children went outdoors in the winter more frequently when educators and the director had OP training, regardless of the number of loose parts present. However, in ELCC centres where educators and directors had less OP training, increased access to loose parts was associated with more frequent OP during winter. This interaction highlights how OP training improves OP frequency in the winter months, and how access to loose parts improves this outcome. Centres where both educators and the director had OP training reported more loose parts on average (5.0 vs. 4.3). A hierarchical model outlining the results with interaction terms included is provided in S3 Appendix.

### Preschool-aged Programs

To understand the influence of the examined explanatory variables on OP opportunities in PS programs, the same final hierarchal regression model used to investigate I/T outcomes was employed for each PS outcome variable (Table 8). Auspice was statistically associated with summer OP duration, with non-profit centres reporting shorter durations of OP on average than for-profit centres (β=-.121). In addition, rural centres exhibited increased summer OP duration (β=.063) and risky play (β=.058). Responding centres with OP-specific training for both directors and educators had longer summer OP duration (β=.081) and increased risky play (β=.092). Educator OP training alone was also significantly associated with summer OP duration (β=.080) for PS programs. As was the case for I/T programs, there was a statistically significant negative effect for PS risky play and more educators with longer tenure (β=-.075). PS programs that provided children or educators with all-weather gear were positively associated with increased risky play (β=.065). Similar to I/T program results, child autonomy was strongly related to all four PS OP outcomes, with the largest effects for winter OP duration (β=.153) and risky play (β=.242).

**Table 8.** Regression model results for ELCC centres serving preschool-aged programs, all outcomes.

|  |  | All Outcomes: PS programs, Betas & Significance |  |  |  |
| --- | --- | --- | --- | --- | --- |
|  |  | Duration Summer | Duration Winter | Frequency Winter | Risky Play |
| Auspice | Non-profit vs. For-profit | **-.121\*\*\*** | -.058 | -.006 | .010 |
| Rural/Urban | Rural vs. Urban Postal Code | **.063\*** | -.026 | -.031 | **.058\*** |
| Centre Ages | Both Ages vs. Just PS | .029 | **.099\*\*** | **-.612\*\*\*** | **.082\*\*** |
| OP Training | Director Only vs. None | .004 | .015 | .011 | .017 |
|  | Educator Only vs. None | **.080\*** | -.003 | -.021 | .045 |
|  | Director and Educators vs. None | **.075\*** | .023 | .032 | **.101\*\*** |
| Staff Tenure | Medium vs. Low | .000 | -.021 | -.022 | **-.075\*** |
|  | High vs. Low | -.002 | -.018 | -.023 | -.035 |
| OP Area Size | Smaller/Off-Site vs. As required | **.168\*\*\*** | -.027 | -.010 | **.093\*** |
|  | Larger vs. As required | .088 | .008 | -.027 | .018 |
|  | Much Larger vs. As Required | **.218\*\*\*** | .075 | .016 | .064 |
| Access to OP Area | Direct vs. Not | .014 | -.054 | -.017 | -.002 |
| Affordances | Loose Parts | **.096\*\*** | **.235\*\*\*** | **.181\*\*\*** | **.249\*\*\*** |
|  | Gardening Elements | .060 | .005 | -.033 | .067 |
|  | Natural Elements | .043 | **.095\*** | **.128\*\*\*** | .074 |
|  | Fixed Equipment | **.106\*\*** | -.015 | -.030 | **.146\*\*\*** |
|  | Portable Equipment | .053 | -.051 | -.022 | -.065 |
| All-Weather Gear | Children or Educators vs. Neither | .039 | .056 | -.004 | **.065\*** |
|  | Children and Educators vs. Neither | -.002 | -.003 | -.009 | .0049 |
| Child Autonomy | Medium vs. Low | **.105\*\*** | **.074\*** | .043 | **.095\*\*** |
|  | High vs. Low | **.104\*\*** | **.153\*\*\*** | **.108\*\*\*** | **.242\*\*\*** |
|  | Adjusted R<sup>2</sup> | .138 | .101 | .451 | .282 |
|  | N (weighted) | 1,003 | 1,013 | 1,010 | 1,017 |

\* $p < .05$ , \*\* $p < .01$ , \*\*\* $p < .001$

PS programs with smaller than required OP spaces or that relied on off-site OP areas to meet licensing requirements had statistically longer OP duration in summer (β=.168), and more risky play (β=.093) than centres that met licensing’s minimum on-site size requirements (but did not exceed). Having much larger OP areas than required by licensing was also significantly associated with increased PS OP duration in the summer (β=.218). Of the five categories of OP affordances, the number of types of loose parts was most strongly related to the PS outcomes, reaching statistical significance for all four outcomes. The number of natural elements was positively associated with winter outcomes, both OP frequency (β=.128) and OP duration (β=.095). The number of types of fixed equipment was positively associated with the amount of risky play (β=.146).

A hierarchical model outlining the results with interaction terms included is provided in S3 Appendix. The largest interaction effect (β=.139) was between director and educator training and child autonomy, predicting winter OP duration. When both educators and directors had OP training, the mean winter OP duration was 2.2 hours per day, compared to 2.0 hours per day when only one or the other was trained.

## Discussion

The MECO Pan-Canadian survey fills an important data gap, representing the first national data examining the existing provision of OP in Canadian ELCC settings. Findings from this study present a current snapshot of OP provision in Canadian ELCC programs, providing a baseline data point from which to assess future changes. These results also examine the factors that are associated with children’s OP and risky play in ELCC programs to inform strategies and policies to enhance OP provision.

The interpretation of these findings suggests some areas of strength within the Canadian ELCC landscape in relation to children’s OP; I/T programs frequently reported daily OP in summer and winter seasons, and the majority of I/T and PS programs reported larger than required OP areas. However, the results from this study also highlight important areas for future attention and consideration, including limited opportunities for risky play and child autonomy within OP provision in Canadian ELCC programs.

Encouragingly, the results from the MECO survey indicate that most Canadian ELCC centres go outdoors every day. However, all types of programs displayed a distinct seasonal trend for time spent outdoors, both in frequency and duration, with less time outdoors in winter than in summer. PS programs showed the most fluctuation in time outdoors across seasons, with an average 1.24 hours per day and 2 days per week less time outdoors in winter than summer. Comparatively, I/T programs reported, on average, similar decreases in OP duration between winter and summer (1.23 hours per day), but demonstrated minimal differences in seasonal OP frequency. ELCC centres serving both I/T and PS programs showed a median of 2 days per week spent outdoors in winter, compared to 5 days per week in ELCC centres serving only PS. This is concerning since mixed-age group centres comprised 75% of our sample. The seasonal effect found in our data aligns with existing research highlighting reduced OP time in seasons that may experience more adverse weather conditions, such as snow, rain, wind and cold temperatures (56,57). Seasonal trends in OP frequency and duration are also indicative of macrosystem level ideologies and approaches towards perceived adverse weather conditions that influence OP participation, as seen in previous studies (22,58–60). Descriptive results surprisingly demonstrated lower mean days outdoors per week in the winter for PS programs (2.9 days/week) than for I/T programs (4.5 days/week). While this warrants further investigation in future studies, we hypothesize that I/T programs may report more frequent time outdoors not necessarily because children are playing outdoors, but because they are being strapped into strollers for outdoor walks.

Risky play was limited among responding ELCC centres, echoing similar research (61,62), and reflecting the macrosystem-level societal concerns for injury and litigation within the ELCC landscape (32,58,63). Play at heights and rough-and-tumble play were the most common, while use fire and play with tools were the least common types of risky play in I/T and PS programs. Overall, PS programs engaged in more risky play than I/T programs, particularly play at heights. Similarly, Sandseter et al.’s Norwegian study reported age as positively associated with total amount of risky play and demonstrated that play at heights was among the most prevalent forms of risky play occurring outdoors in ELCC centres, however rough-and-tumble play was not as prominent in their sample (62). Risk-taking opportunities and challenge are a critical component of child development, and the results from our survey suggest that further supports are needed to expand all types of risky play in ELCC centres.

Below, we outline the main patterns that emerged from the findings, and consider them in the context of the key ingredients for children’s OP of time, space and freedom, and the levels of the Ecological Systems Model – the microsystem, mesosystem, exosystem and macrosystem (31).

### Training

Our findings reinforce the importance of training, particularly ensuring that all centre staff receive training related to OP. Among 58% of the ELCC centres in our sample, either the educators or the director, or both had received OP training (Table 5). The importance of training is well-recognized in helping ELCC professionals to build the skills to integrate time, space and freedom into their practice (32,51). ELCC centres where both the director and educators had OP training demonstrated more frequent OP for I/T programs in winter, and more time spent in OP for PS programs in summer, as well as allowing for more risky play (both types of programs). They also had more loose parts on average in I/T centres and allowed PS children more autonomy. This suggests that training is better integrated into programs when learning happens across all professionals, rather than among a few isolated members of staff. This is further supported by past research in which ELCC educators identified the importance of having a consistent understanding and shared approach with colleagues and the support of senior management to effectively provide OP, and particularly risky play, at their centre (32).

Our results highlight an inverse relationship between a higher proportion of long-tenured educators and lower levels of risky play, further suggesting the importance of training on this particular subject. This finding may reflect the difficulty of shifting entrenched routines and mindsets for ELCC centres with long-standing educators. Because the research on risky play and consequently training to support it are relatively recent developments (12,51), we would expect that longer-serving educators may be less likely to have received this training than those newer to the field.

Child autonomy was among the strongest correlates of all outcomes in I/T and PS aged programs, demonstrating that even for infants, it is important to provide children the right to decide when and how they spend time outdoors. Within our sample, 51% of I/T programs and 44% of PS programs indicated that participating children were never or rarely given the autonomy to decide when to go outdoors. By comparison, only 15% of I/T programs and 18% of PS programs often or always allowed the children to decide (Table 6). Training that incorporates the importance of child autonomy is essential for empowering educators to prioritize children’s freedom and agency in OP. By learning how to respect children’s voices and see them as competent and capable of making their own decisions, educators can provide one of the essential ingredients of OP – *freedom*. This, in turn, allows children to spend more time outdoors and engage in greater levels of risky play. In addition, Tonge et al. (64) found that when children had continuous access to the outdoor environment at any time during the day the quality of the educator-child interaction increased [69]. Free routines that offer children autonomy to enter the outdoor space of their own accord have also been shown to increase physical activity, learning and development, and extended play opportunities (64–66).

Overall, the results highlight the importance of implementing easily accessible and regular training opportunities for all ELCC centre staff to help challenge entrenched practices and align with current OP approaches, including OP in all seasons and the provision of risky play. Quality OP and risky play professional development and training opportunities contribute to enhanced OP opportunities in ELCC environments (microsystem), and support sharing the benefits of OP with others (mesosystem), including families and colleagues (58,67). This training needs to be well integrated into post-secondary programs, as well as readily available as ongoing professional development opportunities (68).

### OP Provision: All-weather Gear, Affordances, and Space

Our findings reveal that ELCC centres with I/T programs that equipped both children and educators with all-weather gear reported longer OP durations in summer and winter, along with increased levels of risky play (Table 7). Among PS centres, this effect was found for winter play duration and risky play (Table 8). In both cases, this was a weak association, but indicates a notable trend, which is also consistent with past research. Weather-related challenges are a commonly reported barrier to OP, particularly in winter (32) and provision of suitable clothing can address this issue, allowing both children and educators to participate in outdoor activities comfortably and safely, regardless of weather conditions.

Consistent with the importance of having a stimulating *space* as a key ingredient of OP, greater play affordances within the ELCC centres, including loose parts and other play elements (natural, gardening elements, and fixed equipment) enhanced OP. In particular, the availability of loose parts was identified as a critical variable strongly associated with extended OP durations in both summer and winter. For I/T centres, loose parts were related to increased OP frequency and duration in winter, particularly when educators lacked OP training. Interestingly, risky play did not show a direct association with loose parts but was instead linked to the variety of gardening elements and fixed equipment. This connection aligns with the common use of these elements for activities such as playing at heights and using tools. Of note, greater counts of gardening elements (planters, garden beds, etc.) significantly increased OP duration in I/T programs in the winter and summer, but were not associated with PS program OP outcomes. Natural elements (trees, shrubs, etc.) were positively associated with OP duration and frequency in PS programs in the winter; however, I/T outcomes were not associated with natural elements. These findings may be due to I/T programs using gardening elements as a structured way to interact with the natural environment.

Overall, the results indicate the importance of microsystem level factors, including available affordances and the provision of all-weather gear, to foster environments that promote longer and more meaningful OP experiences for all children.

### Within and Beyond the Fence: Space Size and Access

The importance of *space* characteristics was reinforced in findings examining the size and accessibility of the licensed outdoor space, as well as the use of off-site “beyond the fence” areas to support OP provision. For I/T programs having an on-site OP area larger or much larger than required by licensing was associated with increased OP duration in summer and OP frequency in winter (Table 7). Having a much larger OP area than required was associated with greater duration of OP in summer for PS programs (Table 8). Direct access to the OP area was associated with increased OP duration in the summer and risky play for I/T programs, however it was not related to any outcomes for PS programs. Existing evidence outlines that OP areas adjacent to the indoor space are associated with enhanced OP opportunities in ELCC programs (69,70).

Although direct on-site access to outdoor space is critical to support OP provision, our findings also suggest there are advantages of also incorporating off-site “beyond the fence” OP areas into regular ELCC programming. PS centres that also utilized off-site spaces experienced longer durations of OP in the summer and more opportunities for risky play. These off-site environments can offer different affordances then ELCC on-site outdoor areas, including more expansive spaces, other natural elements, and diverse physical challenges that promote exploration and creativity. Additionally, beyond the fence locations likely include local parks and playgrounds with play equipment for older children, thereby encouraging challenge and risky play opportunities.

### Limitations

The MECO survey offered valuable insights into the current state of OP provision across licensed full-day ELCC centres in Canada. However, there were several limitations which should be acknowledged. The target population for survey respondents was limited to regulated, full-day centre-based ELCCs, excluding nursery schools, licensed family child care, and programs providing only part-day or school-age care. While this allowed for a shared context for survey respondents, this exclusion limited the generalizability of findings to all child care programs in Canada. Future research could expand the sample to include part-day programs and family child care programs, providing a broader understanding of OP practices across different types of ELCC environments.

Additionally, our sample was not representative of the ELCC landscape with respect to our overall response rate (9.8%), as well as from different regions and auspices. The survey’s promotion strategy encountered challenges impacting respondent reach, particularly in Alberta and Ontario, where direct distribution through government contacts was not feasible. Likewise, we were not provided permission to distribute the survey in the NWT. This resulted in inconsistent survey respondent representation from this region and impacted our territory area sample frame. This speaks to the challenge of conducting a national survey across a vast geographic region with heterogeneity in early childhood administration. Further, 30% of our sample was from the for-profit sector, in contrast to 50% across Canada (71). It is likely that the for-profit ELCC centres in our sample were not representative of the population, influencing the generalizability of the results. Future directions include exploring robust engagement strategies, such as partnering early with provincial regulatory bodies or leveraging alternative distribution networks, to ensure comprehensive coverage across all regions. Additionally, mobilizing provincial and territorial academic institutions, agencies or organizations to conduct regional surveys may lead to more comprehensive national representation.

Automated bot responses during data collection for the MECO survey presented a significant challenge. Initial survey promotion included various social media platforms, resulting in a significant influx of suspicious, non-human responses. Measures were put in place to identify and exclude bot responses.However, it is possible that some fraudulent responses were inadvertently included or, conversely, valid responses were excluded during the validation process. In addition, the necessary halt to social media promotion may have limited our engagement reach, in particular among provinces where a distribution list was not obtainable. Future surveys could implement advanced verification techniques such as real-time automated response pattern monitoring. Additionally, altering the promotion channels could minimize the risk of bot responses, including limiting social media promotion and focussing efforts on contacting trusted child care organizations.

The inherent limitations of survey research mean that causal inferences cannot be drawn from these findings. The MECO survey was designed to collect baseline data that illustrate patterns of association across various OP outcomes. These findings provide valuable insights into current conditions and correlations, offering a descriptive overview to guide future inquiry. This baseline serves as a critical first step, laying the groundwork for tracking change over time and supporting the development of more rigorous, causal research in the future.

## Conclusion and Future Directions

The MECO survey provides insights into the current status of OP in ELCC centres at a timely moment in the history of child care in Canada. The vast changes currently happening in the Canadian ELCC landscape, with significant federal and regional investments in child care provision, are resulting in considerable increases in the availability and affordability of child care for families in Canada. OP has not been a focus during these expansions, yet there is increasing interest and recognition by many sectors of its importance. By recognizing how time, space, and freedom for OP interact to support children’s OP opportunities, policymakers, ELCC practitioners and advocates can develop comprehensive strategies to promote change at multiple levels. A holistic approach is required that addresses the complex interplay between individual beliefs, institutional resources, policies, and societal values in creating supportive environments for OP. These strategies can include advocating for policy changes that prioritize OP at the macrosystem and exosystem levels, raising societal awareness of OP importance, including among families and educators within the microsystem centre level, and ensuring children are empowered to make their own decisions about their OP.

The findings from the MECO survey highlight opportunities for critical policy recommendations to support ELCC programs in Canada. First, the provision of all-weather gear for children and educators as a standard to support OP in all seasons. Second, access to quality educational training on outdoor and risky play both in pre-service (post-secondary) and post-service (ongoing professional development) for educators. Third, encouraging ELCC centres to continually monitor and enhance their OP spaces and associated affordances, including the inclusion of loose parts and natural features. Relatedly, the wealth of existing research on best-practice outdoor space design for children’s play (38,72) should be conveyed in accessible formats to ELCC operators. Fourth, awareness on the importance of outdoor and risky play must be further communicated to families, educators and others to shift societal views away from safety, litigation and weather fear-based approaches to OP provision (73,74).

There is the potential to conduct the MECO survey at regular intervals to monitor how future initiatives influence OP provision, providing data-driven insights to inform decisions. Regular data collection can provide benchmarks to foster meaningful progress towards furthering the ingredients of time, space and freedom in children’s OP provision in ELCC environments.

## Authorship contribution statement

RR conceptualized the study, developed the research questions, oversaw data collection, and contributed to the manuscript; BF conceptualized the study, conducted statistical analyses, and edited the manuscript; HH assisted with data collection and contributed to the manuscript; CH assisted with data collection, wrote the first draft and edited the manuscript; MB, CB, JC, MM and all members of the advisory committee (see Acknowledgements) provided oversite of the study, assisted with distribution of the survey; and edited or reviewed the manuscript; MB conceptualized the study, developed the research questions, obtained funding, supervised statistical analyses and led writing of the manuscript. All authors have read and approved the final manuscript.

## Data availability

All dataset files will be available from the UBC Research Data Collection database following acceptance.

## Acknowledgments

We respectfully acknowledge the traditional territories and lands of the First Nations, Inuit and Métis people upon which our work has been conducted and compiled. We thank the early childhood educators that participated in the survey and the insightful advisory committee that guided this important work. We also thank the invaluable contribution of this project’s advisory committee, including Marc Battle, Jane Beach, Caroline Bouchard, Jeff Crane, Beverlie Dietze, Megan McPhee, Sylvie Melsbach, Michal Perlman, Christina Pickles and Cathy Poole, who supported this project. Thank you to the Lawson Foundation, the Lyle S. Hallman Foundation and the Muttart Foundation for supporting this work. We also extend our gratitude to Christine Alden from the Lawson Foundation for providing feedback on this manuscript.

## References

1. Tremblay M, Gray C, Babcock S, Barnes J, Bradstreet C, Carr D, et al. Position Statement on Active Outdoor Play. Int J Environ Res Public Health. 2015 Jun 8;12(6):6475–505.

2. Dankiw KA, Tsiros MD, Baldock KL, Kumar S. The impacts of unstructured nature play on health in early childhood development: A systematic review. Peña Garay M de L, editor. PLoS One [Internet]. 2020 Feb 13 [cited 2020 Feb 17];15(2):e0229006.

3. Mann J, Gray T, Truong S, Brymer E, Passy R, Ho S, et al. Getting Out of the Classroom and Into Nature: A Systematic Review of Nature-Specific Outdoor Learning on School Children’s Learning and Development. Front Public Health [Internet]. 2022 May 16 [cited 2024 Apr 7];10:877058.

4. Ernst J, Burcak F. Young children’s contributions to sustainability: The influence of nature play on curiosity, executive function skills, creative thinking, and resilience. Sustainability [Internet]. 2019 Aug 4 [cited 2019 Aug 22];11(15):4212. =

5. Hinkley T, Brown H, Carson V, Teychenne M. Cross sectional associations of screen time and outdoor play with social skills in preschool children. PLoS One. 2018;13(4).

6. Brussoni M. Outdoor risky play. In: Burns T, Gottschalk F, editors. Education in the Digital Age: Healthy and Happy Children [Internet]. Paris: Organization for Economic Cooperation and Development; 2020. p. 53–68.

7. Gray C, Gibbons R, Larouche R, Sandseter EBH, Bienenstock A, Brussoni M, et al. What is the relationship between outdoor time and physical activity, sedentary behaviour, and physical fitness in children? A systematic review. Int J Environ Res Public Health. 2015;12(6).

8. Truelove S, Bruijns BA, Vanderloo LM, O’Brien KT, Johnson AM, Tucker P. Physical activity and sedentary time during childcare outdoor play sessions: A systematic review and meta-analysis. Vol. 108, Preventive Medicine. Academic Press Inc.; 2018. p. 74–85.

9. Lund Fasting M, Høyem J. Freedom, joy and wonder as existential categories of childhood– reflections on experiences and memories of outdoor play. Journal of Adventure Education and Outdoor Learning. 2024;2:145–58.

10. Liang J, Pu Y, Chen J, Liu M, Ouyang B, Jin Z, et al. Global prevalence, trend and projection of myopia in children and adolescents from 1990 to 2050: a comprehensive systematic review and meta-analysis. Br J Ophthalmol [Internet]. 2025 [cited 2025 Apr 1];109:362–71.

11. Zhang X, Chen Y, Jin S, Bi X, Chen D, Zhang D, et al. Association of serum 25-Hydroxyvitamin D with Vitamin D intervention and outdoor activity among children in North China: an observational study. BMC Pediatr [Internet]. 2020 Dec 1 [cited 2025 Apr 24];20(1):542.

12. Brussoni M, Gibbons R, Gray C, Ishikawa T, Sandseter EBH, Bienenstock A, et al. What is the relationship between risky outdoor play and health in children? A systematic review. Int J Environ Res Public Health. 2015;12(6):6423–54.

13. Sandseter EBH. Characteristics of risky play. Journal of Adventure Education & Outdoor Learning. 2009 Jun 24;9(1):3–21.

14. Kleppe R, Melhuish E, Sandseter EBH. Identifying and characterizing risky play in the age one-to-three years. European Early Childhood Education Research Journal. 2017;25(3).

15. Sandseter EBH, Kleppe R, Kennair LEO. Risky play in children’s emotion regulation, social functioning, and physical health: an evolutionary approach. Int J Play [Internet]. 2023 Dec 19 [cited 2022 Dec 20];12(1):127–39.

16. Sandseter EBH, Kennair LEO. Children’s risky play from an evolutionary perspective: The anti-phobic effects of thrilling experiences. Evolutionary Psychology. 2011;9(2):257–84.

17. Kleppe R, Sandseter EBH, Sando OJ, Brussoni M. Children’s dynamic risk management – a comprehensive approach to children’s risk willingness, risk assessment, and risk handling. Int J Play [Internet]. 2024 Nov 19 [cited 2024 Nov 18];1–15.

18. Dodd HF, Lester KJ. Adventurous play as a mechanism for reducing risk for childhood anxiety: A conceptual model. Clin Child Fam Psychol Rev [Internet]. 2021 Jan 19 [cited 2021 Jan 21];24:164–81.

19. Gray P. Risky Play: Why Children love and need it. In: Loebach J, Little S, Cox A, Owens PE, editors. The Routledge Handbook of Designing Public Spaces for Young People: Processes, Practices and Policies for Youth Inclusion. 1st ed. London: Routledge; 2020. p. 39–51.

20. Mullan K. A child’s day: trends in time use in the UK from 1975 to 2015. British Journal of Sociology [Internet]. 2019 Apr 11 [cited 2019 Jun 6];70(3):997–1024.

21. Boxberger K, Reimers A, Boxberger K, Reimers AK. Parental correlates of outdoor play in boys and girls aged 0 to 12—A systematic review. Int J Environ Res Public Health [Internet]. 2019 Jan 11 [cited 2019 Jan 30];16(2):190.

22. Lee EY, Bains A, Hunter S, Ament A, Brazo-Sayavera J, Carson V, et al. Systematic review of the correlates of outdoor play and time among children aged 3-12 years. International Journal of Behavioral Nutrition and Physical Activity [Internet]. 2021 Dec 18 [cited 2021 Mar 31];18(1):1–46.

23. Lambert A, Vlaar J, Herrington S, Brussoni M. What is the relationship between the neighbourhood built environment and time spent in outdoor play? A systematic review. Int J Environ Res Public Health [Internet]. 2019 Oct 11 [cited 2019 Nov 25];16(20):3840.

24. Holt NL, Neely KC, Spence JC, Carson V, Pynn SR, Boyd KA, et al. An intergenerational study of perceptions of changes in active free play among families from rural areas of Western Canada. BMC Public Health [Internet]. 2016 Aug 19 [cited 2023 Jul 18];16(1):1–9.

25. Russell W, Barclay M, Tawil B. Playing and being well: A review of recent research into children’s play, social policy and practice, with a focus on Wales [Internet]. Cardiff: PlayWales; 2024 [cited 2024 Nov 23].

26. Herrington S, Nicholls J. Outdoor play spaces in Canada: The safety dance of standards as policy. Crit Soc Policy. 2007 Feb 1;27(1):128–38.

27. Brunelle S, Herrington S, Coghlan R, Brussoni M. Play worth remembering: Are playgrounds too safe? Child Youth Environ. 2016;26(1):17.

28. Brussoni M, Brunelle S, Pike I, Sandseter EBH, Herrington S, Turner H, et al. Can child injury prevention include healthy risk promotion? Injury Prevention. 2015 Dec 22;21:344–7.

29. Organization for Economic Cooperation and Development. PF3.2: Enrolment in childcare and pre-school [Internet]. Paris; 2024 [cited 2024 Nov 23].

30. Statistics Canada. Child care arrangements for children 0 to 5 years, 2023 [Internet]. 2023 [cited 2025 Jan 30]. Available from: https://www150.statcan.gc.ca/n1/pub/11-627-m/11-627-m2023070-eng.htm

31. Bronfenbrenner U. Ecology of the family as a context for human development: Research perspectives. Dev Psychol. 1986;22(6):723–42.

32. Cheng T, Brussoni M, Han C, Munday F, Zeni M. Perceived challenges of early childhood educators in promoting unstructured outdoor play: an ecological systems perspective. Early Years [Internet]. 2023 Feb 26 [cited 2022 Feb 27];4–5:904–20.

33. Leggett N, Newman L. Play: Challenging educators’ beliefs about play in the indoor and outdoor environment. Australasian Journal of Early Childhood. 2017;42(1):24–32.

34. Dietze B, Kashin D. Perceptions that early learning teachers have about outdoor play and nature. Learn Landsc. 2019;12:91–105.

35. Wyver S, Tranter P, Naughton G, Little H, Sandseter EBH, Bundy A. Ten ways to restrict children’s freedom to play: The problem of surplus safety. Contemporary Issues in Early Childhood. 2010;11(3):263–77.

36. Kernan M. Space and place as a source of belonging and participation in urban environments: Considering the role of early childhood education and care settings. European Early Childhood Education Research Journal. 2010 Jun;18(2):199–213.

37. Akbari E, McCuaig K, Mehta S. Early Childhood Education Report 2023 [Internet]. Toronto; 2024 [cited 2025 Jan 30].

38. Herrington S, Lesmeister C. The design of landscapes at child-care centres: Seven Cs. Landsc Res. 2006 Jan 23;31(1):63–82.

39. Herrington S, Brunelle S, Brussoni M. Outdoor play spaces in Canada: As if children mattered. In: Waller T, Ärlemalm-Hagsér E, Sandseter EBH, Lee-Hammond L, Lekies K, Wyver S, editors. The SAGE handbook of outdoor play and learning. London: SAGE; 2017. p. 143–65.

40. Lawson Foundation. More than a new course: A framework for embedding outdoor and land-based pedagogies in post-secondary ECE programs [Internet]. Toronto; 2024 Jun [cited 2024 Nov 23].

41. Ball DJ, Brussoni M, Gill TR, Harbottle H, Spiegal B. Avoiding a dystopian future for children’s play. Int J Play. 2019;8(1):3–10.

42. Alden C, Pyle A. Multi-sector perspectives on outdoor play in Canada. Int J Play. 2019 Sep 2;8(3):239–54.

43. Oberle E, Zeni M, Munday F, Brussoni M. Support factors and barriers for outdoor learning in elementary schools: A systemic perspective. Am J Health Educ [Internet]. 2021 Aug 4 [cited 2021 Aug 3];1–15.

44. Smedsrud TM, Kleppe R, Lenes R, Moser T. Early Childhood Teachers’ Support of Children’s Play in Nature-Based Outdoor Spaces—A Systematic Review. Educ Sci (Basel) [Internet]. 2024 Jan 1 [cited 2025 Feb 20];14(1):13.

45. Beaulieu E, Beno S. Healthy childhood development through outdoor risky play: Navigating the balance with injury prevention. Paediatr Child Health [Internet]. 2024 Jul 22 [cited 2024 Jul 23];29(4):255–61.

46. Pasolli L, Child Care Now. An analysis of the Multilateral Early Learning and Child Care Framework and the Early Learning and Child Care Bilateral Agreements [Internet]. Ottawa; 2019 Nov [cited 2024 Nov 23].

47. Department of Finance Canada. Government of Canada. 2022 [cited 2024 Nov 26]. Supporting early learning and child care.

48. Government of Canada. 2021 Canada country background report on early learning and child care: Background [Internet]. 2021 [cited 2024 Nov 26].

49. Taylor A, Bourassa L, Liadsky B, Ellard-Gray A. The power of a relational values-driven approach to building adult capacity: Final evaluation report of Outdoor Play Strategy 2.0. Toronto; 2024 Jun.

50. McCuaig K, Bertrand J. Policy oversight of outdoor play in Early Childhood Education Setting in Canadian Provinces and Territories [Internet]. Toronto; 2019 [cited 2024 Nov 26].

51. Lawson Foundation. Advancing outdoor play and early childhood education: A discussion paper [Internet]. Toronto; 2019 [cited 2024 Nov 23].

52. Sando OJ, Sandseter EBH. Affordances for physical activity and well-being in the ECEC outdoor environment. J Environ Psychol [Internet]. 2020 Apr 30 [cited 2020 May 3];101430.

53. Harris PA, Taylor R, Thielke R, Payne J, Gonzalez N, Conde JG. Research electronic data capture (REDCap)—a metadata-driven methodology and workflow process for providing translational research informatics support. J Biomed Inform. 2009;42(2):377–81.

54. Friendly M, Feltham L, Mohamed SS, Nguyen NT, Vickerson R, Forer B. Early childhood education and care in Canada 2019. 12thEdition ed. Toronto: Childcare Resource and Research; 2021. 1–279 p.

55. Spencer RA, Joshi N, Branje K, Lee McIsaac J, Cawley J, Rehman L, et al. Educator perceptions on the benefits and challenges of loose parts play in the outdoor environments of childcare centres. AIMS Public Health [Internet]. 2019 [cited 2019 Nov 3];6(4):461–76

56. Jones J, Wolfenden L, Grady A, Finch M, Bolsewicz K, Wedesweiler T, et al. Implementation of continuous free play schedules in Australian childcare services: A cross-sectional study. Health Promotion Journal of Australia [Internet]. 2020 Apr 1 [cited 2025 Jan 30];31(2):199–206.

57. Predy M, Holt N, Carson V. Examining correlates of outdoor play in childcare centres. Canadian Journal of Public Health [Internet]. 2021 Apr 1 [cited 2021 Mar 31];112(2):292–303.

58. Richard B, Turner J, Stone MR, McIsaac JLD. How an early learning and child care program embraced outdoor play: A case study. Journal of Childhood, Education and Society. 2023 Oct 10;4(3):306–21.

59. Sandseter EBH, Cordovil R, Hagen TL, Lopes F. Barriers for outdoor play in Early Childhood Education and Care (ECEC) institutions: Perception of risk in children’s play among European parents and ECEC practitioners. Child Care in Practice. 2020 Nov 22;26(2):111–29.

60. MacQuarrie M, McIsaac JLD, Cawley J, Kirk SFL, Kolen AM, Rehman L, et al. Exploring parents’ perceptions of preschoolers’ risky outdoor play using a socio-ecological lens. 10.1080/1350293X20222055103 [Internet]. 2022 Mar 23 [cited 2022 Apr 7];1–16.

61. Loebach J, Ramsden R, Cox A, Joyce K, Brussoni M. Running the risk: The social, behavioral and environmental associations with positive risk in children’s play activities in outdoor playspaces. Journal of Outdoor and Environmental Education [Internet]. 2023 Dec 1 [cited 2024 Dec 19];26(3):307–39.

62. Sandseter EBH, Kleppe R, Sando OJ. The prevalence of risky play in young children’s indoor and outdoor free play. Early Child Educ J [Internet]. 2021 Jun 25 [cited 2020 Jul 2];49:303–12.

63. Sandseter EBH, Sando OJ. “We Don’t Allow Children to Climb Trees”: How a Focus on Safety Affects Norwegian Children’s Play in Early-Childhood Education and Care Settings. Am J Play. 2016;8(2):178–200.

64. Tonge KL, Jones RA, Okely AD. Quality interactions in early childhood education and care center outdoor environments. Early Child Educ J. 2019;47:31–41.

65. Hesketh KR, van Sluijs EMF. Features of the UK childcare environment and associations with preschooler’s in-care physical activity. Prev Med Rep. 2016 Jun 1;3:53–7.

66. Siraj-Blatchford I. Conceptualising progression in the pedagogy of play and sustained shared thinking in early childhood education: A Vygotskian perspective. Educational and Child Psychology. 2009;26(2):77–89.

67. Spencer RA, Joshi N, Branje K, Murray N, Kirk SF, Stone MR. Early childhood educator perceptions of risky play in an outdoor loose parts intervention. AIMS Public Health [Internet]. 2021 [cited 2021 Mar 7];8(2):213–28.

68. Dietze B, Cutler A. College faculty’s outdoor play pedagogy: The ripple effect. Canadian Journal of Environmental Education. 2020;23(2):86–105.

69. Herrington S, Lesmeister C, Nicholls J, Stefiuk K. Seven C’s: An informational guide to young children’s outdoor play spaces. Vancouver: Consortium for Health, Intervention, Learning and Development (CHILD); 2007.

70. Ceppi G, Zini M. Children, Spaces, Relations: Metaproject for an Environment for Young Children. Reggio Emilia, Italy: Reggio Children; 1998.

71. Friendly M, Beach J, Gayaththiri A, Cossette A, Lillace J, Forer B. Early Childhood Education and Care in Canada 2023 [Internet]. Toronto; 2024 Aug [cited 2024 Nov 28].

72. Morgenthaler T, Lynch H, Loebach J, Pentland D, Schulze C. Using the Theory of Affordances to Understand Environment–Play Transactions: Environmental Taxonomy of Outdoor Play Space Features—A Scoping Review. American Journal of Occupational Therapy [Internet]. 2024 Jul 1 [cited 2024 Oct 6];78(4).

73. Brussoni M, Han CS, Lin Y, Jacob J, Munday F, Zeni M, et al. Evaluation of the web-based OutsidePlay-ECE intervention to influence early childhood educators’ attitudes and supportive behaviors toward outdoor play: Randomized controlled trial. J Med Internet Res [Internet]. 2022 Jun 10 [cited 2022 Jun 9];24(6):e36826.

74. Brussoni M, Han CS, Lin Y, Jacob J, Pike I, Bundy A, et al. A web-based and in-person risk reframing intervention to influence mothers’ tolerance for, and parenting practices associated with children’s outdoor risky play: Results of a randomized controlled trial. J Med Internet Res. 2021;23(4):e24861.

